# Michigan men’s diabetes project II: protocol for peer-led diabetes self-management education and long-term support in Black men

**DOI:** 10.1101/2022.11.03.22281893

**Authors:** Jaclynn Hawkins, Srijani Sengupta, Katherine A. Kloss, Katie Kurnick, Alana Ewen, Robin Nwawkwo, Martha Funnell, Jamie Mitchell, Lennette Jones, Gretchen Piatt

## Abstract

Previous literature has indicated that Black men are twice as likely to develop type 2 diabetes compared to their non-Hispanic White counterparts and are also more likely to have associated complications. Furthermore, Black men have lower access to quality health care, and masculinity norms have been shown to hinder them from seeking the limited care that is available. In this study, we aim to investigate the effect of peer-led diabetes self-management education and long-term ongoing support on glycemic management. The first phase of our study will consist of modification of existing diabetes education content to be more appropriate for the population of interest, Then, in the second phase, we will conduct a randomized controlled trial to test the intervention. Participants randomized to the intervention arm will receive diabetes self-management education, structured diabetes self-management support, and a more flexible ongoing support period. Participants randomized to the control arm will receive diabetes self-management education. Diabetes self-management education will be taught by certified diabetes care and education specialists, while the diabetes self-management support and ongoing support period will be facilitated by fellow Black men with diabetes who will be trained in group facilitation, patient-provider communication strategies, and empowerment techniques. The third phase of this study will consist of post-intervention interviews and dissemination of findings to the academic community. The primary goal of our study is to determine whether long-term peer-led support groups in conjunction with diabetes self-management education are a promising solution to improve self-management behaviors and decrease A1C levels. We will also evaluate the retention of participants throughout the study, which has historically been an issue in clinical studies focused on the Black male population. Finally, the results from this trial will determine whether we can proceed to a fully-powered R01 trial or if other modifications of the intervention are necessary.

**Trial Registration:** Registered at ClinicalTrials.gov with an ID of NCT05370781 on May 12, 2022

## INTRODUCTION

### Background and Rationale

In the United States, more than 37.3 million Americans, or 11.3% of the population, are living with diabetes [1]. Disparities in diabetes prevalence and complications also exist among racial/ethnic groups [2]. For instance, Black people are twice as likely to have type 2 diabetes (T2D) compared to non-Hispanic White people [2]. Black men have worsened glycemic management (e.g. checking blood glucose levels daily) compared to non-Hispanic White men, their risk for T2D complications is higher and they are more likely to die earlier from these complications [2, 3].

Extant research in health disparities shows how poverty and race can influence the health and wellbeing trajectories of Black men [2]. For instance, T2D has its most damaging effects within urban low-income communities of color, and yet they continue to lack access to quality health care [2]. While low-income Black men have similar rates of T2D compared to Black women, Black men are disproportionately impacted with respect to severity and mortality [2]. In addition to being less likely to follow diabetes self-management plans and engage in diabetes self-management behaviors (e.g. checking blood glucose levels daily), low-income Black men face a multitude of additional challenges to maintaining a healthy diet, such as limited availability of healthy foods [2-5]. Black men also often experience barriers that interfere with diabetes management behaviors such as a lack of social support, negative patient-provider relationships, cost and long work hours [6, 7]. Though existing studies find that Black men are at elevated risk for suboptimal diabetes management, they are also less likely to participate in research studies that test diabetes self-management interventions [8]. Additionally, Black male participants are more likely to drop out of diabetes self-management intervention trials before their completion, resulting in additional problems [8].

There is a new emphasis in recent research on the critical role of gender in the management of health behaviors. This body of work highlights how male gender norms can conflict with both healthy practices and engagement in healthcare services [2, 9]. For instance, among men, the need to exhibit toughness, confidence, suppress emotions while exuding independence and control can pose a barrier to engagement with care (i.e. following treatment plans) [9]. Existing studies also reveal how male gender norms can impede utilization of critical social support offered by family and community networks, including the acceptance of compassionate environments (i.e. emotional support) [9].

Peer leaders (PLs) are trained lay individuals used to advance diabetes self-management in minority communities through encouraging and helping with goal setting, problem solving, and social, emotional, and peer support [10-13]. They are chosen based on their cultural background and community connections. The use of PLs is an essential component of interventions aimed at improving diabetes health outcomes among low-income Black communities [10-13]. Yet, despite the documented effectiveness of peer-led diabetes interventions on bettering diabetes-related health behaviors and health outcomes, this model has largely not been adapted to meet the needs of Black men. It is also important to note that among studies of peer leader models, participants and peers are largely women [8]. An opportunity exists to tailor messaging and intervention content to better serve Black men living with type 2 diabetes.

While there is an absence of research that utilizes Black male peer leaders (i.e., male lay helpers) in support of Black men with diabetes, male lay helpers have provided Black men with health education and emotional support in other areas of health care; including interventions for the prevention of chronic illness, such as for HIV and hypertension [14, 15]. Gender-specific programming utilizing male leaders to reduce stigma related to health care seeking has also been recommended by Black men [16]. More studies are needed to establish the efficacy of these approaches and to determine whether the use of male PLs can improve health outcomes among Black men with diabetes.

Research also suggests that tailoring diabetes self-management education and support (DSME/S) interventions to address the needs of Black men can play an instrumental role in improving their health outcomes [7, 17]. The National Standards for Diabetes Care and the National Standards for Diabetes Self-Management Education and Support highlight the relevance of providing initial DSME and on-going DSMS that is individualized to assist people with diabetes in maintaining effective self-management, aiming to improve health equity across different populations [18]. DSMS interventions, including those delivered by professionals and peer leaders, bettered A1C and self-management outcomes in comparison to control groups in previous studies [19-22]. Piatt et al. studied the use of a 12 session, 15-month intervention that combined DSME directed by a diabetes educator with DSMS delivered by peer leaders (PL) in 9 Black community churches in metro-Detroit [23]. The intervention completion rate was high (96.6%) [23]. Twelve peer leaders (n = 12) were trained in how to facilitate goal setting, skills development, and group cohesion [23]. After the professional DSME was provided, 6 monthly DSMS groups were facilitated by 12 PL (mean age: 61.9 years, 100% Black, 25% male), followed by a supplemental 6 months of further support to assess the logistical feasibility of sustaining DSMS efforts [23].

As previously stated, there is a paucity of diabetes intervention research that focuses on the unique needs of Black men. Peer-led education and support interventions have been successful, but Black men have been poorly recruited and retained in these studies [8]. Discourses on gender equity in health often focus on a binary between men and women, which often ignore the structural and systemic issues that sub-groups are required to navigate. This limited framing of gender equity does not acknowledge the complex health and social inequities faced by marginalized groups of men, particularly those relating to race, age, socioeconomic status, geography and disability. Therefore, a more concerted and nuanced focus on health equity in the field of men’s health is needed. In the context of our proposed project, we seek to create T2D programming that addresses the intersection of gender and markers of marginalization (e.g. race/ethnicity; income). It is our expectation to provide evidence for the effectiveness of ongoing peer-led DSME/S intervention for Black men with T2D. The findings will facilitate understanding of barriers and facilitators to implementing this community-based approach and the impact this initiative has on the individual and community levels.

We propose to tailor an existing peer intervention by 1) using male peers as interventionists and 2) modifying the intervention content to focus on messaging appropriate for men. Our study will then assess the effectiveness of the adapted, peer-led DSMS and ongoing support period. Success of this study sets the stage for larger implementation trials and adoption of evidence-based interventions to address foundations of health disparities for Black men living with type 2 diabetes.

### Objectives

This study’s objectives are twofold: 1) in collaboration with Black men with type 2 diabetes, we will adapt an evidence-based peer leader intervention designed to improve diabetes self-management to better fit the needs of this population; and 2) conduct a pilot randomized controlled trial (RCT) to evaluate participant recruitment and retention rates, treatment and intervention satisfaction and estimate intervention effect sizes on our primary outcomes self-management behaviors and glycemic control measured through hemoglobin A1C (A1C) as well as on secondary outcomes such as diabetes social support at baseline, 3-, 9- and 15-months. Our previous study, Michigan Men’s Diabetes Project (MenD), measured health outcomes for Black men with T2D who participated in peer-led support group interventions (Hawkins et al, 2022, not published). Based on findings from MenD, the present study (hereafter referred to as MenD II) will be modified to include a 6-month period of group based ongoing support in addition to an added emphasis on patient-provider communication strategies.

## MATERIALS AND METHODS

### Study Design

This proposed study, MenD II, includes a developmental phase (adjustment of the intervention, including stakeholder feedback, followed by feasibility testing with Black men), a validation phase (RCT), and a dissemination phase. The dissemination phase includes: post-intervention interviews/focus groups, and sharing of findings with academics and community members.

#### Phase I: Intervention Adaptation

In this study, we will adapt the existing intervention from previous peer leader research and the MenD study for Black men with type 2 diabetes to add additional components, with the goal to better meet the needs of Black men living with diabetes [23]. During Phase I we will conduct 14 interviews to better understand factors that impact the implementation of a peer-led diabetes self-management education support (PLDSMS) intervention adapted for Black men with T2D in the US. Interviewees will be diabetes educators, community partners, participants from previous peer leader studies, peer leaders from previous studies, and researchers in the fields of diabetes and/or men’s health and physical health. These individuals will be familiar with working with lay health workers, experienced with our target population, and/or knowledgeable about the research topic. Phase I of this project was approved by the University of Michigan Health Sciences and Behavioral Sciences Institutional Review Board (HUM00200496) on June 6, 2021. The interviews in phase I will be guided by the Tailored Implementation in Chronic Disease (TICD) framework, a comprehensive framework describing the determinants of implementation success [24]. Determinants of practice are the barriers and facilitators that might impact on implementation of an intervention. The TICD Checklist includes 57 potential determinants of practice grouped into seven domains [24]. These seven domains include guideline factors, individual health professional factors, patient factors, professional interactions, incentives and resources, capacity for organizational change, and social, political and legal factors [24]. The aim of the interviews is to understand the structural, contextual and cultural factors impacting the implementation of a peer-led diabetes education and support intervention adapted for Black men with T2D. We will use data from these interviews to inform the implementation of the intervention in Phase II, a tailored PLDSMS RCT. At this point, participant recruitment and educational materials will be adapted for gender. It is essential that intervention materials are adapted to account for language, culture, health literacy and urban poverty contexts.

Standardized treatment goals and the DSMS protocol will be adapted to include local references, cultural values and familiar situations. Though many changes cannot be predicted, based on the existing literature, our study team will develop a DSMS module that will emphasize teaching male participants how to engage in clinic appointments with their diabetes and other health care providers (i.e. sharing concerns, being assertive to advocate for needs, and asking questions).

#### Phase II: RCT

During Phase II, an RCT will be conducted of the modified intervention to measure participant recruitment and retention rates, treatment and intervention satisfaction and estimate intervention effect sizes on our primary outcomes (i.e., A1C and self-management behaviors) as well as on secondary outcomes (i.e., diabetes social support and diabetes-related distress). Phase II of this study was approved by the University of Michigan Medical School Institutional Review Board (HUM00200469) on November 11, 2021.

The RCT will be conducted among N = 64 Black male adult residents of southeastern Michigan. Four men will be recruited and trained to function as peer leaders, with sixty men randomized to a control group or to the tailored PLDSMS (See Fig 1). Our study team hypothesizes that 1) participants in the adapted PLDSMS approach will have overall improved outcomes over the control group, and 2) participants in the PLDSMS will learn diabetes self-management skills at greater levels than participants in the control group.

**Fig 1.**
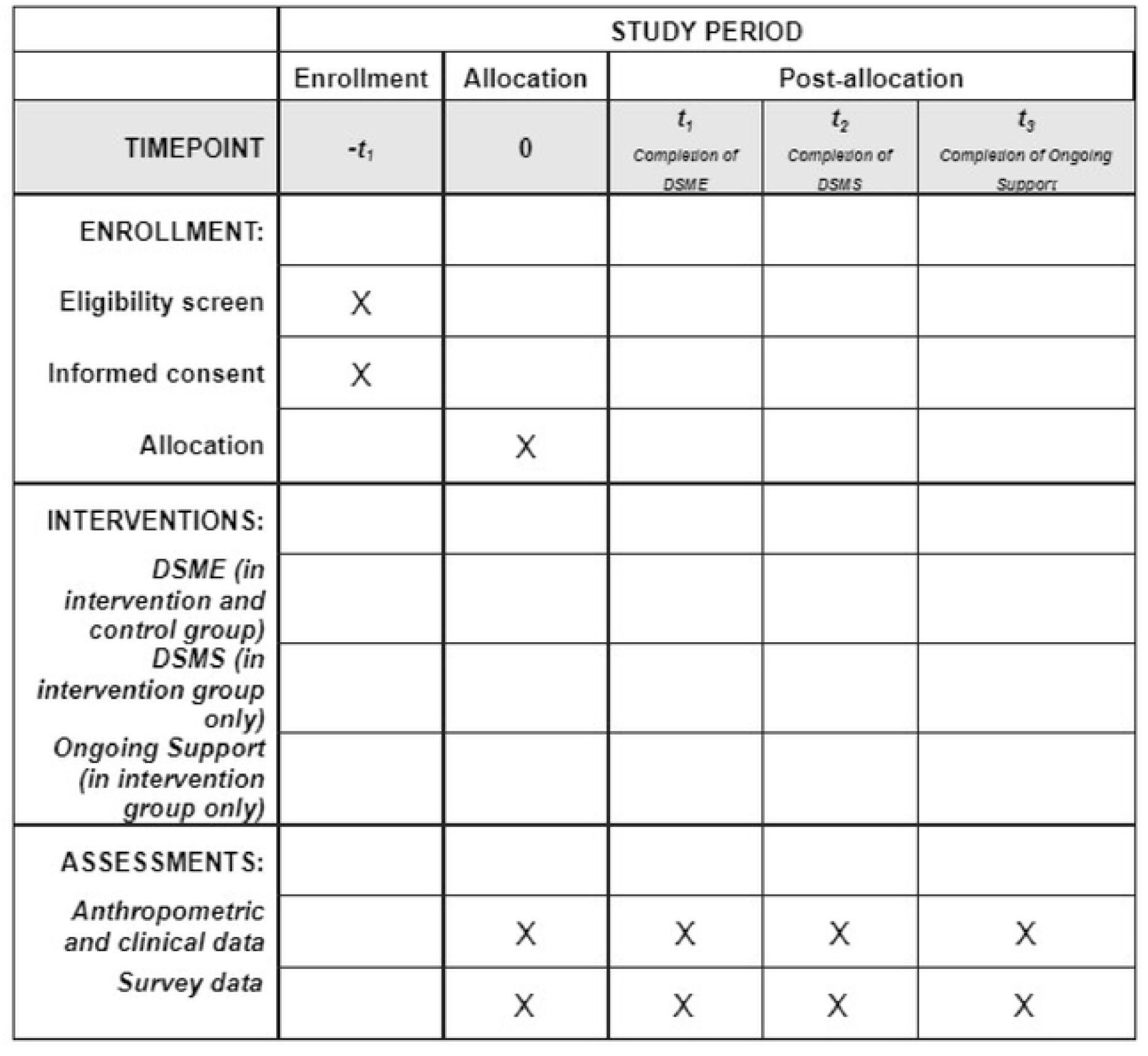
RCT Enrollment, Intervention, and Assessments

To measure growth, the primary outcomes will be change in A1C levels and self-management behaviors [25-27]. Secondary outcomes measured will include BMI, blood pressure, quality of life, depressive symptoms, empowerment, social support, barriers and resources, and adherence to gender norms [28-34]. Primary and secondary outcomes will be measured at the baseline visit and at each follow-up visit.

#### Phase III: Dissemination

Phase III will consist of three parts; administering post-intervention interviews and focus groups with both participants and stakeholders, statistical analysis, and dissemination of findings. Activities to share results will include presentation of findings to academic and community forums. In academia, research findings will be presented to the annual meeting of the American Diabetes Association, with the preparation of at least 3 manuscripts. To share results in the community, community-based dissemination of findings will include a report and presentation that will be shared electronically with researchers, community-supported, translational health research that are focused on recruiting and working with African American men in diabetes research. A toolkit will be provided to a local senior center in southeast Michigan and will contain community resources, and data collection tools to allow for continuation of learning, and functions staff need to play to sustain improvements in outcomes.

### Study Setting

Due to the risks associated with COVID-19, all DSME/S and ongoing support sessions will be held virtually via a Health Insurance Portability and Accountability Act (HIPAA) compliant virtual platform, Zoom for Health at U-M [35]. Post-treatment and follow-up self-report questionnaires will be completed in person, telephonically, or virtually. All biometrics will be collected in person at a local church or at a local senior center. To facilitate privacy, confidentiality, and safety in each location, rooms with doors, and telephones, will be available. In person health assessments will follow up to date local, state, and national COVID-19 protocols and recommendations. The data for this study will be collected at baseline, 3 months (DSME completion), 6 months (DSMS completion), and 15 months (ongoing support completion).

### Eligibility criteria

#### Inclusion criteria

To be eligible for the study, participants (n=64) will be Black/African American men ages 21 years or older who have had a diagnosis of type 2 diabetes for a six-month period or longer. Previous studies done by this team focused on older Black men who were at least 55. The range of participants has been expanded in order to increase recruitment and allow analysis of age differences in our sample population. Additionally, it is essential that participants have transportation to attend program activities, be under the care of a physician addressing their diabetes, and be willing to attend both group delivered virtual sessions and in-person health assessments.

Four of the participants will serve as peer leaders and will attend 30 hours of training to learn skills needed to facilitate DSMS. In addition to the general inclusion criteria listed for participants, the peer leaders must also have at least an 8th grade education, have had type 2 diabetes for over one year, be actively working on their own self-management goals, and be ready to go through the training and be a peer leader.

This study team contemplated restricting eligibility to a higher-risk population of participants with A1C ≥ 8%. A majority of the preliminary data suggest that over 50% of the proposed study sample will have an A1C ≥ 8%. By focusing on all Black men with diabetes, it allows us to cast a wide net for secondary prevention and public health impact. Potential participants who meet eligibility criteria will be invited to partake in the baseline screening assessment. While the above eligibility criteria have been chosen based on previous studies, adjustments will be made to the future, larger trial, based on results and feedback from our proposed study.

#### Exclusion criteria

The exclusion criteria for this study includes 1) non-ambulatory, serious health conditions (i.e., severe symptomatic heart disease, visual impairment, renal failure, and peripheral neuropathy); 2) psychiatric illness (i.e., severity requiring hospitalization); and 3) serious diabetes complications (i.e., blindness) that would interfere with meaningful participation in the groups.

### Intervention

During Phase II of this study, an 18-month randomized control trial (RCT) will take place and include: 1) diabetes self-management virtual education and support (DSME/S) with peer leaders to assess participant recruitment and retention rates, treatment and intervention satisfaction and estimate trial effect sizes on our primary outcomes of self-management behaviors; and 2) the estimate on secondary outcomes such as glycemic control (A1C), diabetes social support, diabetes-related distress, and adherence to gender norms. To measure outcomes, these data will be collected at baseline, 3-months (DSME completion), 6-months (DSMS completion), and 15-months (ongoing support completion) at a senior center in southeast Michigan, a church in southeast Michigan, telephonically, or virtually. In the Phase II RCT, we will individually randomize enrolled participants using a 50/50 randomization scheme to either the peer led DSMS or a control group. All DSME/S sessions will be held virtually on Zoom. At the time this protocol was submitted, three peer leaders were recruited, enrolled and completed training and participant recruitment had begun.

#### Intervention (peer leader DSMS n = 30)

The participants randomized to the peer-led DSMS group will receive 10-hours of virtual diabetes self-management education (DSME) over the course of 3 months. The 10-hours was chosen to align with what is reimbursable through Medicare for initial diabetes self-management training [36]. A certified diabetes care and education specialists (CDCES) will deliver DSME with two peer leaders co-facilitating one 15-person group. A CDCES facilitating DSME classes ensures consistent and accurate delivery of the diabetes content while covering all required and necessary self-management topics. Research shows that Black men with diabetes experience significant barriers to healthcare, so we are offering DSME with the assumption that most participants have not received formal DSME.

These participants will then transition into six 90-min monthly virtual PLDSMS sessions tailored to Black men with T2D. PLs will facilitate DSMS with the oversight of the CDCES. Though the CDCES will not be present in the PLDMS, the participants will meet with the CDCES once each month to answer questions and get support for the next DSMS session(s) as needed. Additionally, the CDCES will be available via telephone to answer any clinical questions the PLs need support with. In previous studies, we observed that PLs were most effective and confident when they had ongoing support and assistance to uphold their efforts in the areas of clinical content, educational methods, group facilitation, and communication skills. The patient-directed, DSMS session content is standardized around 6 core processes: 1) reflecting on relevant self-management experiences, 2) discussing emotions, 3) problem-solving barriers to diabetes management, 4) addressing questions about diabetes, 5) setting behavioral goals and 6) discussing patient provider communication strategies [23]. During DSME, participants will also be given a guidebook titled, “Diabetes Management Guidebook,” which is culturally specific and literacy-appropriate. The guidebook was initially developed for previous projects and was well received [37].

Upon the completion of DSMS, participants in the PLDSMS group will begin a 6-month period of ongoing support. In this stage, participants will be encouraged to foster ongoing DSMS through programs and initiatives that are meaningful to them. On-going support will build on the peer-led component of this study and encourage participants to engage in the activities of their choice (i.e. forming a walking group, discussing self-management topics, cooking classes, etc.) to improve health outcomes. Peer leaders will not be compensated for this period to assess the logistical feasibility of sustaining DSMS efforts after the study is complete.

#### Control group (Control n = 30)

Participants randomized to the control group will engage in 10 hours of group-delivered virtual DSME provided by a CDCES. These participants will not receive any DSMS or ongoing support from PLs and the PLs will not engage in the DSME sessions. The main purpose of the trial includes evaluation of the recruitment and retention rates, intervention satisfaction, and estimated intervention effect sizes. Taking this into consideration, the enhanced usual care condition was chosen as the control in order to 1) ensure that any intervention effects are not due to provision of diabetes education alone, 2) minimize ethical concerns regarding assignment of underserved populations to receive a no-treatment control and 3) control for improvements due to attention and positive regard and expectancies for improvement due to participation in treatment (i.e., Hawthorne effects).

#### Peer leaders (n = 4)

Candidates recruited to be PLs will participate in PL training (detailed below), co-facilitate DSME sessions with a CDCES, lead DSMS sessions, and complete the same health assessments as all other participants in the study. After the peer leader training is complete, quarterly leader review sessions will be held with the PLs and CDCES so PLs can support one another, ask questions, and further hone group facilitation skills. The number of these sessions will be based on the needs of the PLs and determined in collaboration with the CDCES and the PLs. PLs will be offered $10/h for the training, leader review sessions, DSME sessions, and DSMS sessions. PLs will not be paid during the final ongoing support period to simulate the real-world scenario of facilitating support after the research study has concluded.

### Outcomes

#### Primary outcome measures

Primary outcomes for this study include glycemic control and regimen adherence (See Table 1). The primary measurement used to determine metabolic control will be A1C. We will use a DCA Vantage point-of-care testing instrument to measure A1C. This analyzer is capable of measuring hemoglobin A1C with non-fasting fingerstick in 6 minutes. To measure regimen adherence and self-management behaviors, the Perceived Diabetes Self-Management Scale, the Adherence to Refills and Medicines Scale for Diabetes (ARMS-D), and the Diabetes Care Profile will be utilized [25-27].

**Table 1.**
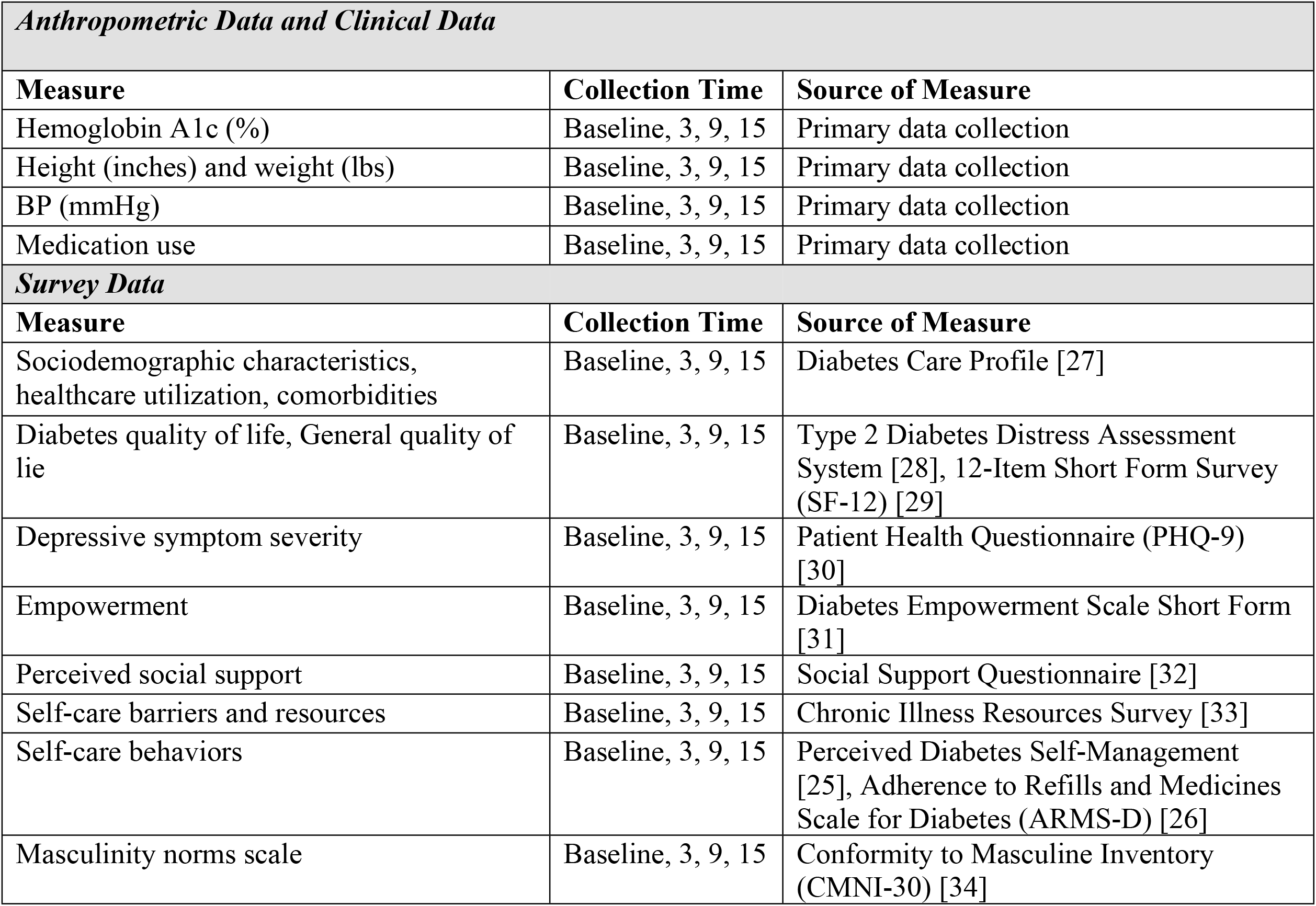
Primary and Secondary Outcome Measures.

| ***Anthropometric Data and Clinical Data*** |  |  |
| --- | --- | --- |
| **Measure** | **Collection Time** | **Source of Measure** |
| Hemoglobin A1c (%) | Baseline, 3, 9, 15 | Primary data collection |
| Height (inches) and weight (lbs) | Baseline, 3, 9, 15 | Primary data collection |
| BP (mmHg) | Baseline, 3, 9, 15 | Primary data collection |
| Medication use | Baseline, 3, 9, 15 | Primary data collection |
| ***Survey Data*** |  |  |
| **Measure** | **Collection Time** | **Source of Measure** |
| Sociodemographic characteristics, healthcare utilization, comorbidities | Baseline, 3, 9, 15 | Diabetes Care Profile [27] |
| Diabetes quality of life, General quality of life | Baseline, 3, 9, 15 | Type 2 Diabetes Distress Assessment System [28], 12-Item Short Form Survey (SF-12) [29] |
| Depressive symptom severity | Baseline, 3, 9, 15 | Patient Health Questionnaire (PHQ-9) [30] |
| Empowerment | Baseline, 3, 9, 15 | Diabetes Empowerment Scale Short Form [31] |
| Perceived social support | Baseline, 3, 9, 15 | Social Support Questionnaire [32] |
| Self-care barriers and resources | Baseline, 3, 9, 15 | Chronic Illness Resources Survey [33] |
| Self-care behaviors | Baseline, 3, 9, 15 | Perceived Diabetes Self-Management [25], Adherence to Refills and Medicines Scale for Diabetes (ARMS-D) [26] |
| Masculinity norms scale | Baseline, 3, 9, 15 | Conformity to Masculine Inventory (CMNI-30) [34] |

#### Secondary outcome measures

To measure adherence to gender norms, the Conformity to Masculine Inventory (CMNI-30) will be utilized [34]. BMI will be calculated using height and weight, with height being measured using a stadiometer and weight being measured using a calibrated digital scale. Blood pressure will be measured 3 times using the oscillometric technique and a final blood pressure will be calculated by taking the average of these measures.

Perceived social support will be measured using the Social Support questionnaire [32], depressive symptom severity will be measured using the Patient Health Questionnaire (PHQ-9) [30], the Type 2 Diabetes Distress Assessment System [28] will be utilized to determine diabetes-related distress and general quality of life will be measured with the 12-Item Short Form Survey (SF-12) [29]. Empowerment will be measured using the Diabetes Empowerment Scale Short Form [31]. The Chronic Illness Resources Survey will be completed to indicate barriers and resources to self-care [33]. As an additional measurement, participants will complete questionnaires that assess socio-demographic, behavioral, psychosocial, and openness to health services utilization and are supported in diverse populations with diabetes (See Table 1).

### Participant Timeline

For this study, data will be collected at baseline, completion of DSME (3-months), completion of DSMS (9-months), and completion of ongoing support period (15-months) (See Table 1).

### Sample Size

#### Power analysis

While we plan to recruit 64 Black men to be our participants for this study, we expect a final sample size of 48 participants (excluding the four participants who will serve as peer leaders), assuming a 20% attrition rate. There will be 12 participants per group, with 2 groups in the control arm and 2 groups in the PLDSMS arm. Assuming correlations of 0.25 between successive measurements of A1C, this sample size will yield a power of 0.8 to detect a difference of 0.6 standard deviations between average values of A1C in the treatment and control groups.

### Recruitment

#### Peer leader recruitment, training, and assessment

Peer leaders will be recruited from the interviews in Phase I, from participants in previous studies who indicated interest in becoming a PL, or by staff at the senior center. After the peer leaders are selected, they will receive about 30 hours of training in facilitation skills, coping strategies, and empowerment-based communication skills. The training sessions will be spread over 3 months in order to minimize burden on the PLs, and they will be compensated at $10/hr. The research staff will work with the PLs to ensure the timing of the training sessions is suitable for all parties. The curriculum and materials for the training will be based on Piatt et al. due to the 100% attendance rate of the 15 PLs in the study [23]. The content will be modified to include specific concerns voiced by Phase I interviewees. The training process will model alternative perspectives that allow for healthy behaviors to be framed as competence and strength, rather than as a challenge to their masculinity.

Peer leaders will be trained in a group setting, and the curriculum will include both didactic learning and skill building with a specific focus on providing support to adults with type 2 diabetes. The training will be led by two CDCESs who helped develop and implement similar training curricula in previous projects [19]. During the training, the PLs will have many opportunities to practice their group facilitation skills. The CDCES and fellow peer leaders will provide constructive and specific feedback to support the PL in enhancing their skill set.

At the conclusion of the training period, the PLs will be given written assessments and a process-based evaluation to assess their understanding of the concepts and skills taught. Post training measures will also be in place to further assess goal-setting, communication, and facilitation abilities.

We will hold quarterly meetings with all the peer leaders to offer additional opportunities to hone skills, share experiences, and offer support to one another. The peer leaders will be encouraged to exchange information and support each other outside these sessions as well.

At the time this protocol was submitted, peer leader recruitment and training had been completed.

#### Participant recruitment

We plan to take a multifaceted approach to participant recruitment. Our first strategy is to utilize paper and electronic advertisements at community partner sites. We will hang flyers describing the study at the senior center and church where biometric data will be collected. These two places will be sent electronic copies of the flyer and they may choose to include it in newsletters. Additionally, the Healthier Black Elder Center will include a recruitment posting in their newsletter and email listings [38].

The second strategy is to use two research registries to reach out to potential participants that meet our inclusion criteria. The first is the Michigan Center for Urban African American Aging Research Participant Research Pool (MCUAAAR PRP) through the Healthier Black Elder Center, a volunteer registry of Black males who are at least 55 years old that is accessible to researchers to find candidates for their studies. Our previous studies have successfully utilized MCUAAAR PRP as a recruitment tool. There are currently a total of 1080 men on the registry, with 339 of them known to have type 2 diabetes [39]. The second registry is the University of Michigan Data Office for Clinical and Translational Research DataDirect PHI system [40]. Both of these registries will provide our research team with names, phone numbers, and email addresses of men who meet our inclusion criteria. Men from these registries will be emailed and/or called to gauge interest and to sign up for enrollment.

Our third strategy is snowball sampling by providing the flyer to a variety of different people including participants of a previous DSMES study, participants who were interviewed during the Phase I of this study, and other researchers. Those who are given the flyer will be encouraged to share it with people in their personal and professional networks who may benefit from the study.

Men interested in participating in the study will either call a central office phone number or be identified through cold calls and/or recruitment emails. If the men express interest in the study, they will be screened to ensure they meet the inclusion criteria previously stated. Upon completion of the phone screen, the study team member will inform the individual if they are eligible to complete the baseline interview. If the individual is eligible, a baseline assessment will be scheduled, and details will be given to the individual.

At the time this protocol was submitted, participant recruitment had begun but no participants were enrolled.

#### Retention

Our team has had high recruitment and retention rates among Black men in our target recruitment region in previous studies. A former diabetes-focused qualitative research study conducted by our team retained 30 out of 32 men. Specific procedures based on our previous work will be implemented to minimize participant attrition. To increase retention: 1) Non-biometric data collection sessions will be completed telephonically or virtually in order to maximize the convenience of data collection for each participant, and 2) Advanced scheduling, multiple reminder letters, and phone reminders will be utilized to encourage participants to attend their in-person appointments.

### Allocation

Participants will be randomized to the peer led diabetes self-management support (PLDSMS) group or a control group using a 50/50 randomization scheme. A computer will create a random number sequence, and then the randomization feature in Research Electronic Data Capture (REDCap) will be used to assign participants [41].

After an individual provides informed written consent at the baseline visit, a study coordinator without prior knowledge of the sequence will randomize the participant via REDCap, allocate the individual to the selected group, and inform the participant of their assigned group.

### Blinding

Blinding is not possible for this study because the nature of the intervention requires active participation in each phase. Participants will be notified of their assigned group and given earned incentives accordingly.

### Data Collection Procedures

All research staff will be trained in standardized measurement and data collection techniques. Quantitative and qualitative assessments will occur for all participants at baseline (0-months), completion of DSME (3-months), completion of DSMS (9-months), and completion of ongoing support period (15-months). All participants, including peer leaders will be invited to a 60-minute appointment at a local senior center or church in the metro-Detroit area for physiological testing. Self-report questionnaires will be completed telephonically or virtually, depending on each participant’s preference. After completing each assessment, participants will be compensated $50. Participants who withdraw from the study will still be invited to participate in data collection appointments at each timepoint. All data will be stored in REDCap, a HIPAA-compliant, web-based application provided by the University of Michigan [41], and in a Microsoft Access database [42].

### Data Management

Participants will be assigned a study identification number after they enroll which will be used on all materials and data for the study’s remainder. Confidentiality is of utmost importance, and our participants will be assured of their anonymity. Participants will not be named in any reports regarding the study data and the data collected will only be used for research purposes. The principal investigator will retain control of all data collected, including questionnaires, audiotapes, and transcribed notes. Data will be stored in a locked file drawer in a locked office, and only the research team will have access.

### Statistical Methods

#### Phase I and Phase III

Qualitative analysis will be conducted on the interviews and focus group discussions from Phases I and III. Interviews and discussions will be recorded and transcribed, and we will develop codes utilizing a grounded theory approach. The research team will formulate a coding manual and definitions based on the text which will guide ongoing coding. Pairs of coders will read the transcripts and codes that achieve 80% agreement on code application will be kept. Analyses will be done using Atlas.ti. The findings from Phase I will be used to further refine intervention content as needed. Post-intervention interviews and focus group discussions from Phase III will be evaluated to determine the effectiveness of the intervention and to identify any domain-specific issues.

#### Phase II

With a conservative estimate of a 20% overall attrition rate, our final sample would be 48 participants (excluding peer leaders), given our initial target sample size of 64 Black men. This will result in 24 participants in each group, which is similar to other pilot intervention trials. One goal of our intervention is to have a low dropout rate. Our past studies have retained 90-95% of participants, and for this study, an intervention retention rate of 75% in the PLDSMS arm will be considered sufficient to proceed to the R01 [23, 43]. There will be no interim data analysis. We plan to use the adapted PLDSMS intervention to power a future R01 based on a minimally clinical important difference (i.e., 0.5% decrease in A1C) rather than based on specific effect sizes from this trial. If we find that the PLDSMS intervention resulted in a small effect on the primary and secondary endpoints, this would suggest that further refinement is needed before proceeding to a fully powered trial.

Our goal is to disseminate findings from the current study and detect between group differences. We will assess statistical significance for the intervention group by time interaction using an Individually Randomized Group-Treatment Trial, with intervention group as a between-subjects factor (2 levels), repeated measurements over time as a within subjects factor (3 levels), a within-subject correlation of .5, and an α of .05, and a non-sphericity correction of .75. The effect size used in this design is Cohen’s f (f = σmeans/σ). The effect of the intervention on diabetes management and A1C (primary outcomes) will be assessed using mixed-effects models. Diabetes management and A1C at baseline (0 months), completion of DSME (3 months), completion of DSMS (9 months), and completion of ongoing support period (15 months) will be used as the dependent variables. Independent variables include intervention group, time of assessment, and interaction between time and intervention group. To take into account correlation between observations, random intercepts and slopes will be included into the model. In the analysis we will follow intention-to-treat principles. Analyses will be conducted with adjusting for stratifying variables (i.e. age group).

### Formal Committee

A Data Safety and Monitoring Board (DSMB) will be established to protect the safety of all participants, and to ensure the integrity of the data collected. We will follow techniques suggested in previous literature to establish this board, which will consist of the Principal Investigator (PI), research team members, and five other people who are not involved with the study [43]. These outside members will all be University professors with proficiency in clinical diabetes intervention research, and they will serve as voting members of the board. The DSMB will hold at least three conference calls over the award period for approximately two hours per call to discuss the progress of the intervention and review research results, if applicable. At the first meeting, the board will elect a chair of the DSMB. To facilitate these conference calls, the principal investigator will prepare a report on the progress of the project to date. This report will be circulated well in advance of the conference call to allow all members ample time to read it. These calls will be scheduled and organized by the study coordinator and will be held at a time convenient for all members.

### Safety/Harms

Participants will be asked to provide finger stick capillary blood samples during assessments to measure A1C levels so that we can assess changes throughout the study. These blood draws have risks similar to any other routine blood sample collection including: minor discomfort, minor pain, bruising, and bleeding at the puncture site. Skilled personnel will perform all blood draws and sample collection to mitigate associated risks.

Participants will also be asked to complete a series of psychosocial questionnaires at each assessment. All of the questionnaires are standardized measures that have been used in our own trials and in other diabetes research. There are no significant risks anticipated related to their completion. However, answering questions in the PHQ-9, the demographic questionnaire, the Diabetes Quality of Life measure, the SF-12, and others may cause minor discomfort for participants [25-34]. In order to alleviate this, breaks will be given to reduce fatigue, and research assistants will be trained to obtain personal information in a sensitive fashion. All participants will be informed that they may discontinue completion of individual items, questionnaires, or the study protocol at any time. Participants who experience significant discomfort will have the option of meeting with the PI. Although prompting patients to review their diabetes care practices and providing them with feedback about their health may cause some emotional discomfort or anxiety, this discomfort could prime patients and their primary care physician to address any problems identified, thus ultimately benefiting the participants.

Another risk of our study is breach of confidentiality of data. All research staff will be trained in research ethics, confidentiality protection, and HIPAA prior to and throughout the study period through the Human Research Protection Program (HRPP) in the University of Michigan Office of Research (UMOR) [44]. Peer leaders will also be trained in the protection of participant confidentiality and must pass standardized testing before interacting with the other participants. Participants will be assigned a study identification number during enrollment that will be used in all study materials and data. Identifiable information will be kept separately from the data and laboratory values. The master list containing participants’ names and study identification numbers will be kept in a locked filing cabinet in the School of Social Work accessible only to the research team. Audio/visual recordings of interviews and group discussions will be stored securely on the University of Michigan Dropbox [45], a HIPAA compliant data storage system, and will only be accessible to study personnel. After the study is completed, these recordings will promptly be deleted. If a participant’s A1C is over 14%, systolic blood pressure value is 200 mm Hg or greater, and/or diastolic blood pressure value is 100 mm Hg or greater, both the patient and their provider will be alerted as this poses a risk to the patient and requires immediate attention. All other reports will not identify individual participants. No persons from the community-based locations utilized in the study will have access to personal health information or participant survey data.

For the purposes of this study, adverse events will be considered any undesirable sign, symptom, or medical condition occurring during the study, whether or not related to the intervention. Adverse events include new events not present during the training period or events that were present during the training period but increased in severity over time. All members of the research team will be trained on identifying an adverse event and instructed to report them to the principal investigator immediately. Additionally, study participants will be provided with the phone number of the project coordinator to report any adverse events that occur during the duration of the study. Each adverse event will be recorded and assessed for its date of onset, duration, severity, seriousness, and relationship to study treatment, and any action/treatment that is required. All adverse events will be collected, analyzed, and monitored using an adverse event form. Our team will verify appropriate reporting of adverse events throughout the study.

### Auditing

To ensure PLs have the skills to facilitate DSMS, the CDCES that was involved in the PL training and also facilitated the DSME classes, will meet with the PLs after each DSME session. During this time the CDCES and PLs will review a skills assessment frequently used during the PL training to assess the PLs skills during that session. This assessment will also cover effective and ineffective interactions during the DSME session to support the PL in increasing their group facilitation skills and confidence before they start DSMS sessions. The DSMS and ongoing support periods are intended to provide a comfortable, confidential environment where the participants can talk to their peers alone. However, to ensure treatment fidelity, three DSMS sessions will be selected at random and recorded and evaluated by our research team.

## DISCUSSION

Successful management of diabetes requires both diabetes education and social support that is appropriate for each patient’s lifestyle and environment [2]. The current set up of our healthcare system is unable to provide Black men living with diabetes the resources and support they need [2]. This project aims to investigate whether the use of gender-matched lay helpers in diabetes self-management education and long-term support is a promising solution to improve the health of this high risk population.

Although literature on community-based diabetes interventions with lay helpers is available, these studies have predominantly focused on non-hispanic white, middle-class populations, and among the studies that have focused on Black populations, women dominate as participants and peer-leaders [8]. The results of these interventions have been compelling, which indicates that a similar use of PLDSMS with Black men may be beneficial. Black men who have participated in previous intervention studies have also voiced that they would be most comfortable with Black males as their lay helpers [16]. Our study will utilize lay helpers during DSME and DSMS who match participants in both gender and race in order to encourage participation and retention throughout the intervention. The intervention proposed also includes an ongoing support period where peer leaders are not compensated and can choose their method of interacting with participants to assess the real-world feasibility of long-term DSMS. We believe using Black male peers as leaders and adapting the intervention content has the potential to improve diabetes outcomes by reinforcing proper self-management behaviors.

The data from this study will enhance diabetes intervention literature by contributing the experience of Black men in an urban setting using an interdisciplinary treatment approach. We will also identify strategies for increasing Black male participation and retention in research studies, as this has proven a challenge in previous literature [8]. Lastly, this pilot RCT will allow us to refine recruitment strategies, training materials, and the implementation protocol to be used in a larger cluster RCT. Data from this study will be disseminated to academics and the sample population community to sustain improvements in diabetes outcomes and advance health equity.

## Data Availability

Deidentified research data will be made publicly available when the study is completed and published.

## Acknowledgements

We would like to thank the University of Michigan School of Social Work, the National Institute of Diabetes and Digestive and Kidney Disease (NIDDK), The Michigan Center for Urban African American Aging Research (MCUAAAR), University of Michigan Data Office for Clinical and Translational Research, and REDCap for supporting this project. We would also like to thank our community partners (St. Patrick Senior Center in Detroit and the Ypsilanti Seventh-day Adventist Church in Ypsilanti) and participants, without them this work would not be possible.

## Ethics

This study protocol outlined in Figure 1 has been reviewed and approved by the University of Michigan Health Sciences Center Institutional Review Board (Phase I: HUM00200496, Phase II and III: HUM00200469). All individuals interested in participation will be required to provide a written informed consent document approved by the University of Michigan Institutional Review Board (UM IRB). At the baseline screening assessment session, eligible participants will complete the Informed Consent form. Consent forms will include all required elements of informed consent, including purpose of the study, duration, voluntary participation, alternatives and right to withdraw. Participants will be told that they will be compensated for each study assessment and that the intervention will be provided to them at no cost. In addition, the consent form will discuss the fact that participants have an equal chance of being randomized to either treatment condition. Participants will be provided with a copy of the informed consent form for their records.

## Data Availability Statement

This manuscript does not report data and the data availability policy is not applicable to our article.

## REFERENCES

1. American Diabetes Association [Internet]. 2022 July 28 [cited 2022 Oct 10]. Arlington: The American Diabetes Association. Available from: https://diabetes.org/about-us/statistics/about-diabetes.

2. Hawkins J. Type 2 diabetes self-management in non-Hispanic Black men: A current state of the literature. Curr Diab Rep. 2019;19: 10. doi: 10.1007/s11892-019-1131-8.

3. Liburd LC, Namageyo-Funa A, & Jack L. Understanding “masculinity” and the challenges of managing type-2 diabetes among African-American men. J Natl Med Assoc. 2007;99: 550–552, 554–558.

4. Lee LT, Jung SE, Bowen PG, Clay OJ, Locher JL, Cherrington AL. Understanding the dietary habits of black men with diabetes. J Nurse Pract. 2019;15: 365–369. doi: 10.1016/j.nurpra.2018.12.023.

5. Thorpe RJ, Kennedy-Hendricks A, Griffith DM, Bruce MA, Coa K, Bell CN, et al. Race, social and environmental conditions, and health behaviors in men. Fam Community Health. 2015;38: 297–306. doi: 10.1097/FCH.0000000000000078.

6. Hawkins J, Mitchell J, Piatt G, Ellis D. Older African American Men’s perspectives on factors that influence type 2 diabetes (T2D) self management and peer-led interventions. Geriatrics. 2018;3: 38–47. doi: 10.3390/geriatrics3030038.

7. Crabtree K, Sherrer N, Rushton T, Willig AL, Agne AA, Shelton T RN, et al. Diabetes connect: African American men’s preferences for a community-based diabetes management program. Diabetes Educ. 2015; 41: 118–126.

8. Sherman L, Hawkins J, Bonner T. An analysis of the recruitment and participation of African American men in type 2 diabetes self-management research: A review of the published literature. Soc Work Public Health. 2016;32: 38–48. doi: 10.1080/19371918.2016.1188742.

9. Hawkins J, Watkins DC, Kieffer E, Spencer MS, Nicklett EJ, Piatt G, et al. An exploratory study of gender identity and its influence on health behavior among African American and Latino men with type 2 diabetes. Am J Mens Health. 2016;11: 344–356. doi: 10.1177/1557988316681125.

10. Heisler M. Overview of peer support models to improve diabetes self management and clinical outcomes. Diabetes Spectr. 2007;20: 214–221. doi: 10.2337/diaspect.20.4.214.

11. Heisler M. Different models to mobilize peer support to improve diabetes self management and clinical outcomes: evidence, logistics, evaluation considerations and needs for future research. Fam Pract. 2010;27: 23–32. doi: 10.1093/fampra/cmp003.

12. Tang TS, Funnell MM, Gillard M, Nwankwo R, Heisler M. The development of a pilot training program for peer leaders in diabetes. Diabetes Educ. 2011;37: 67–77.

13. Tang TS, Nwankwo R, Whiten Y, Oney C. training peers to deliver a church based diabetes prevention program. Diabetes Educ. 2012;38: 519–525. doi: 10.1177/0145721712447982.

14. Maulsby C, Millett G, Lindsey K, Kelley R, Johnson K, Montoya D, et al. A systematic review of HIV interventions for black men who have sex with men (MSM). BMC Public Health. 2013;13: 1.

15. Hess PL, Reingold JS, Jones J, Fellman MA, Knowles P, Ravenell JE, et al. Barbershops as hypertension detection, referral, and follow-up centers for black men. Hypertension, 2007;49: 1040–1046. doi: 10.1161/HYPERTENSIONAHA.106.080432.

16. Hurt TR, Seawell AH, O’Connor MC. Developing effective diabetes programming for black men. Glob Qual Nurs Res. 2015;2:233339361561057. doi: 10.1177/2333393615610576.

17. Hawkins J, Kieffer EC, Sinco B, Spencer M, Anderson M, Rosland AM. Does gender influence participation? Predictors of participation in a community health worker diabetes management intervention with African American and Latino adults. Diabetes Educ. 2013;39: 647–654. doi: 10.1177/0145721713492569.

18. Davis J, Fischl AH, Beck J, Browning L, Carter A, Condon JE, et al. 2022 National Standards for Diabetes Self-Management Education and Support. Diabetes Care. 2022:45, 484–494. doi.org/10.2337/dc21-2396.

19. Funnell MM, Tang TS, Anderson RM. From DSME to DSMS: developing empowerment based self management support. Diabetes Spectr. 2007;20: 221–226.doi:10.2337/diaspect.20.4.221.

20. Norris SL, Chowdhury FM, Van Le K, Horsley T, Brownstein JN, Zhang X, et al. Effectiveness of community health Workers in the Care of persons with diabetes. Diabet Med. 2006;23:544–556. doi:10.1111/j.1464-5491.2006.01845.x.

21. Thompson JR, Horton C, Flores C. Advancing diabetes self management in the Mexican-American population: a community health worker model in a primary care setting. Diabetes Educ. 2007;33 (suppl): 159S–65S. doi: 10.1177/0145721707304.

22. Brown HS, Wilson KJ, Pagan JA, Arcari CM, Martinez M, Smith K, et al. Cost effectiveness analysis of a community health worker intervention for low-income hispanic adults with diabetes. Prev Chron Dis. 2012;9:120074.. doi:10.5888/pcd9.120074

23. Koscielniak NJ, Funnell M, Piatt G. Building infrastructure for diabetes self management support in church-based settings—results of a 15-month cluster-randomized controlled trial. Diabetes. 2018;67: 871. doi: 10.2337/db18-871-P.

24. Flottorp SA, Oxman AD, Krause J, Musila NR, Wensing M, Godycki-Cwirko M, et al. (2013). A checklist for identifying determinants of practice: A systematic review and synthesis of frameworks and taxonomies of factors that prevent or enable improvements in healthcare professional practice. Implementation Science. 2013;8: 35. doi:10.1186/1748-5908-8-35.

25. Wallston KA, Rothman RL, Cherrington A. Psychometric properties of the perceived diabetes self-management scale (PDSMS). J Behav Med. 2007;30: 395–401. doi: 10.1007/s10865-007-9110-y.

26. Mayberry LS, Gonzalez JS, Wallston KA, Kripalani S, Osborn CY. The ARMS-D outperforms the SDSCA, but both are reliable, valid, and predict glycemic control. Diabetes Research and Clinical Practice, 2013; 102: 96–104. doi:10.1016/j.diabres.2013.09.010.

27. Fitzgerald JT, Davis WK, Connell CM, Hess GE, Funnell MM, Hiss RG. Development and validation of the diabetes care profile. Evaluation & the Health Professions. 1996;19: 208–230. doi:10.1177/016327879601900205.

28. Polonsky WH, Fisher L, Hessler D, Desai U, King SB, Perez-Nieves M. Toward a more comprehensive understanding of the emotional side of type 2 diabetes: A re-envisioning of the assessment of diabetes distress. Journal of Diabetes and its Complications. 2022;36: 108103. doi:10.1016/j.jdiacomp.2021.108103.

29. Jenkinson C, Layte R. Development and testing of the UK SF-12. J Health Serv Res Policy. 1997;2: 14–18. doi:10.1177/135581969700200105.

30. Kroenke K, Spitzer RL. The PHQ-9: a new depression diagnostic and severity measure. Psychiatr Ann. 2002;32: 509–15. doi:10.3928/0048-5713-20020901-06.

31. Anderson RM, Funnell MM, Fitzgerald JT, Marrero DG. The diabetes empowerment scale. A measure of psychosocial self-efficacy. Diabetes Care. 2000;23: 739–743. doi:10.2337/diacare.23.6.739.

32. Epino HM, Rich ML, Kaigamba F, Hakizamungu M, Socci AR, Bagiruwigize E, Franke MF. Reliability and construct validity of three health-related self-report scales in HIV-positive adults in rural Rwanda. AIDS Care. 2012;24: 1576–1583. doi:10.1080/09540121.2012.661840

33. Glasgow RE, Toobert DJ, Barrera M, Strycker LA. The Chronic Illness Resources Survey: Cross-validation and sensitivity to intervention. Health Education Research. 2005;20: 402–409. doi: 10.1093/her/cyg140

34. Levant RF, McDermott R, Parent MC, Alshabani N, Mahalik JR, Hammer JH. Development and evaluation of a new short form of the Conformity to Masculine Norms Inventory (CMNI-30). Journal of Counseling Psychology 2020;67: 622–636. doi:10.1037/cou0000414.

35. Zoom at U-M [Internet]. (no date) [Cited 2022 Oct 15]. Ann Arbor: Information and Technology Services. Available from: https://its.umich.edu/communication/videoconferencing/zoom.

36. Medicare reimbursement guidelines for DSMT. 2021 Feb 4 [Cited 2022 Oct 15]. Atlanta: Centers for Disease Control and Prevention. Available from: https://www.cdc.gov/diabetes/dsmes-toolkit/reimbursement/medicare.html

37. Tang TS, Gillard ML, Funnell MM, Nwankwo R, Parker E, Spurlock D, Anderson RM. Developing a new generation of ongoing diabetes self-management support interventions. The Diabetes Educator. 2005;31: 91–97. doi: 10.1177/0145721704273231

38. Healthier black elders center(Hbec). (no date) [Cited 2022 Oct 15]. Detroit: Institute of Gerontology. Available from: https://iog.wayne.edu/about/healthier-black-elders-center.

39. Participant resource pool. (no date) [Cited 2022 Oct 15]. Ann Arbor: Michigan Center for Urban African American Aging Research. Available from: https://mcuaaar.org/cores/community-liaison-and-recruitment-core/participant-resource-pool/.

40. Self-serve data tools. (no date) [Cited 2022 Sept 20]. Ann Arbor: Office of Research. Available from: https://research.medicine.umich.edu/our-units/data-office-clinical-translational-research/self-serve-data-tools.

41. Redcap access. (no date) [Cited 2022 Sept 20]. Ann Arbor: Michigan Institute for Clinical and Health Research. Available from: https://michr.umich.edu/redcap-access. CTSA: UL1TR002240

42. Database Software and Applications. (no date) [Cited 20 Sep 2022]. Redmond: Microsoft. Available from: https://www.microsoft.com/en-us/microsoft-365/access

43. Piatt G, Provenzano AM, Nwankwo R, Hall D, Kloss KA, Hawkins JM, Funnell MM. 62-OR: Fostering Sustainability through Diabetes Self-Management Support in African-American Churches: Results of the Praise Diabetes Project. Diabetes. 2021; 70. doi: 10.2337/db21-62-OR.

44. Damocles Study Group. A proposed charter for clinical trial data monitoring committees: Helping them to do their job well. Lancet. 2005;365:711–22. doi: 10.1016/S0140-6736(05)17965-3.

45. Human research protection program (Hrpp). 2022 April 20 [Cited 2022 Sept 20]. Ann Arbor: Research Ethics & Compliance Available from https://research-compliance.umich.edu/human-subjects.

46. Dropbox at U-M. (no date) [Cited 2022 Sept 20]. Ann Arbor: Health Information Technology & Services Availa ble from: https://hits.medicine.umich.edu/collaboration/files-content/dropbox-u-m.

